# Mathematical modeling of SARS-CoV-2 variant substitutions in European countries: Transmission dynamics and epidemiological insights

**DOI:** 10.1101/2023.10.12.23296888

**Authors:** Víctor López de Rioja, Aida Perramon, Sergio Alonso, Cristina Andrés, Andrés Antón, Antoni E. Bordoy, Jordi Càmara, Pere-Joan Cardona, Marti Català, Daniel López, Sara Marti, Elisa Martró, Verónica Saludes, Clara Prats, Enrique Alvarez-Lacalle

## Abstract

**Background:** Countries across Europe have faced similar evolutions of SARS-CoV-2 VOCs, including the Alpha, Delta, and Omicron variants.

**Materials and Methods:** We used data from GISAID and applied a robust, automated mathematical substitution model to study the dynamics of COVID-19 variants across Europe over a period of more than two years, from late 2020 to early 2023. This model identifies variant substitution patterns and distinguishes between residual and dominant behavior. We used weekly sequencing data from 19 European countries to estimate the increase in transmissibility (∆β) between consecutive SARS-CoV-2 variants. In addition, we focused on large countries with separate regional outbreaks and complex scenarios of multiple competing variants.

**Results:** Our model accurately reproduced the observed substitution patterns between the Alpha, Delta, and Omicron major variants. We estimated the daily variant prevalence and calculated ∆β between variants, revealing that: (*i*) ∆β increased progressively from the Alpha to the Omicron variant; (*ii*) ∆*β* showed a high degree of variability within Omicron variants; (*iii*) a higher ∆*β* was associated with a later emergence of the variant within a country; (*iv*) a higher degree of immunization of the population against previous variants was associated with a higher ∆*β* for the Delta variant; (*v*) larger countries exhibited smaller ∆*β*, suggesting regionally diverse outbreaks within the same country; and finally (*vi*) the model reliably captures the dynamics of competing variants, even in complex scenarios.

**Conclusions:** The use of mathematical models allows for the precise and reliable estimation of daily cases of each variant. By quantifying ∆*β*, we have tracked the spread of the different variants across Europe, highlighting a robust increase in transmissibility trend from Alpha to Omicron. On the other hand, we have shown that the country-level increases in transmissibility can always be influenced by the geographical characteristics of the country and the timing of the emergence of the variant.

## 1. Introduction

Since the emergence of the severe acute respiratory syndrome coronavirus 2 (SARS-CoV-2) in late 2019, the virus responsible for the Coronavirus disease 2019 (COVID-19) pandemic has evolved significantly, leading to the emergence of different viral variants responsible for successive waves of infection. To date, over 750 million confirmed cases and more than 6.5 million deaths have been documented worldwide [1]. In particular, several European countries have experienced waves of infection from these variants, highlighting the need for vigilant surveillance and a deep understanding of their transmission dynamics.

Among these variants, some have been designated by the World Health Organization (WHO) as Variants of Concern (VOCs) or Variants of Interest (VOIs) due to their potential impact on transmissibility, disease severity, and the efficacy of diagnostics and vaccines [2], [3]. The VOCs, including Alpha, Beta, Gamma, Delta, and various Omicron variants exhibit distinct characteristics affecting their transmissibility and potential for immune evasion.

The evolution of SARS-CoV-2 has resulted in significant genomic changes that have had a substantial impact on its spread across Europe [4]. Initially dominated by various lineages such as B.1.177, B.1.160, and B.1.258, the introduction of the more transmissible Alpha variant changed the landscape in European countries, leading to an increase in case numbers and hospitalizations [5], [6], [7]. This was followed by the Delta variant, which overtook Alpha and challenged public health efforts due to its high transmissibility, potential for increased disease severity, and partial immune evasion [8], [9], [10]. Subsequently, Omicron and its different subvariants (BA.1, BA.2, BA.5, BQ.1, and XBB) have emerged sequentially, replacing each other with unique substitution patterns [11], [12], [13].

Understanding the dynamics of variant substitution and changes in transmissibility is essential to design effective public health strategies. In this study, we explore these substitution dynamics across Europe using a mathematical model, aiming to unravel similarities and differences between countries. Our model can process large datasets and automatically detect variant behavior, providing a valuable tool for understanding the historical evolution of the pandemic and plausible short-to-mid-term scenarios for a large number of countries. We estimate the daily prevalence of each SARS-CoV-2 variant by country and calculate the increase in transmissibility, Δ*β*, for each substitution and country. We also examine how factors such as initial emergence day and country surface area affect variant behavior. Using corrected daily number of cases, we assess shifts in the effective reproduction number, Rt, associated with emerging variants. Special attention is given to large countries and scenarios where multiple variants are in strong competition, providing nuanced insights into complex dynamics. To our knowledge, while some studies have conducted similar analyses on a large database [14], [15], [16], none have obtained comprehensive results across Europe.

## 2. Materials and Methods

Data on the weekly number of sequenced variants and the number of vaccinated individuals in European countries was obtained from the European Centre for Disease Prevention and Control (ECDC) database. Instead, daily COVID-19 case numbers for each country was obtained from the WHO database1, as ECDC only provides data on a weekly basis. Additionally, we used demographic, geographic, and statistical data obtained directly from the official European statistical website2.

### 2.1. SARS-CoV-2 variants data bases

Our study was based on official variant sequencing data for each European country, obtained from the ECDC website. These data, available on a weekly basis [17], were derived from two primary sources: the Global Initiative on Sharing All Influenza Data (GISAID)3 [18] and the European Surveillance System (TESSy)4. Throughout the COVID-19 pandemic, both databases have provided up-to-date case numbers, variant distributions across Europe, and other essential information such as vaccination rates. Both databases are comprehensively represented in **Suppl. Mat. Tables 1a and 1b**. However, depending on the data source, the number of samples per country and date varied considerably. We found significant inconsistencies suggesting that the epidemiologic surveillance centers reported exclusively to only one source, or data appearing to be duplicated, indicating that some countries report the same sequenced samples to both GISAID and TESSy. Differences in data processing and quality control procedures between the two databases may also contribute to these discrepancies. See **Suppl. Mat. Text S1** for a detailed description of both data sources.

**Table 1.** Results for Europe and for the average of all countries.

|  | Alpha | Delta | Omicron |
| --- | --- | --- | --- |
| Europe | 0.4041 | 0.7821 | 0.9114 |
| Median | 0.4477 | 0.7863 | 1.177 |

Given these issues, we decided to use only GISAID data in the main manuscript and leave the TESSy analysis for the Supplementary Material. There were two main reasons for this decision: GISAID has been the most widely used source in the literature, and it provides data that allowed us to analyze the first significant substitution (*pre-Alpha vs Alpha*), which was not possible with TESSy data, as it provides sequenced variant data primarily since early 2021. By relying exclusively on GISAID, we increased the rigor of our study, albeit at the cost of excluding some countries from our analysis. We explored a combination of the two sources to test the robustness of our results and to provide results for more countries in Suppl. Mat. Text S11.

### 2.2 Data: GISAID database overview

Data from GISAID on variant sequencing begins in early 2020 for most countries, initially presenting a limited number of samples and high variability. Regarding variant assignation, many of these samples are labeled as “Other” in the ECDC database as they predate the definition of VOCs and VOIs by the health authorities at the time of their occurrence5. As mentioned in the Introduction, various lineages circulated in Europe before the Alpha VOC took hold. Among these, B.1.177 was dominant in many countries, with significant proportions of other lineages, such as B.1.160, and B.1.258, also present. These, along with smaller proportions of additional variants, will be referred to hereafter as the *pre-Alpha* package to maintain consistency with the source. As the pandemic progressed across Europe, the number of samples sequenced increased in all countries, with peaks during the waves driven primarily by the Alpha, Delta, and Omicron VOCs. From the period of Alpha dominance to the present, our “Other” package refers to a mix of residual variants found in different countries, not exclusively the *pre-Alpha* package mentioned above, and not exclusively the “Other” category from the GISAID source.

Data from some countries presented issues which did not allow us to conduct the analysis. Some countries, including Hungary, Liechtenstein, and Malta, were excluded from our analysis due to low sampling rates and significant variability in their data. Additionally, we decided not to use the GISAID data from countries with different problems in the datasets such as Austria, Cyprus, Estonia, Greece, Iceland, Luxembourg, Portugal, and Slovakia. Given the different nature of the problems, we refer the reader to the **Suppl. Mat. Text S2** for an explanation of each country. In the end, our study included data from 19 countries: Belgium, Bulgaria, Croatia, Czechia, Denmark, Finland, France, Germany, Ireland, Italy, Latvia, Lithuania, the Netherlands, Norway, Poland, Romania, Slovenia, Spain, and Sweden.

### 2.3 Substitution model and estimation of cases per variant

In this study, we present an enhanced version of a previously used substitution model [19]. Our updated model not only facilitates the analysis of substitution patterns of various SARS-CoV-2 variants across countries but also automatically determines the behavior of each variant during the substitution process. It distinguishes between variants that are major drivers of substitution and those with a marginal presence. Additionally, the model calculates the increase in transmissibility, ∆*β*, for each variant and substitution process. A brief explanation of the model (mathematical and computational) is given below; for a more detailed discussion, refer to **Suppl. Mat. Text S3**.

The simplest scenario for a substitution model involves only two variants: the initially dominant one (1) and the one that will ultimately prevail (2). We assume that both variants evolve according to an exponential dynamic equation 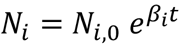, and that the mean time between infection and maximum infectivity of individuals τ is fixed for both variants. Thus, we can relate the exponent *β*_i_ to the transmissibility and thus to the effective reproduction number as 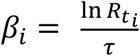. Since τ is fixed, the increase in transmissibility parameter, Δ*β* = *β*_2_ − *β*_1_, remains constant, although *β*_1_ and *β*_2_ may change with time. In this case, we derive the following equation for the fraction of the emerging variant during the substitution period:

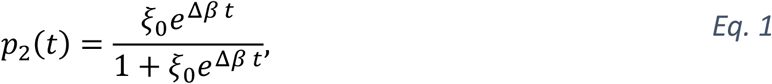

where *ξ*_0_is the initial ratio between variant 2 (future dominant variant) and variant 1 (previous dominant variant). Therefore, for the latter, which is in decline, we find *p*_1_(*t*) = 1 − *p*_2_(*t*).

These equations are useful when we aim to compare Δ*β* between only two variants. However, in general, they may not adequately capture the dynamics of the virus within a country, when multiple variants circulate simultaneously. Results from this simplified approach are presented in **Suppl. Mat. Text S7**, while a comprehensive analysis of all ∆*β* values can be found in the **Suppl. Mat. Text S12**.

When three or more variants are involved, additional equations are included to account for the increased complexity. Assume that *N* variants are competing to dominate the national viral landscape. For each of these competing variants, *i*, we can rewrite the previous Eq. 1 as:

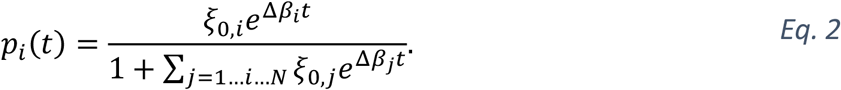

Again, the fraction corresponding to the descending variant can be estimated as p_*desc*_(*t*) = 1 − Σ_*j*=1…*i*…*N*_ *p*_*j*_(*t*).

But, as explained before, not all variants show a peak behavior. Some variants present different trends: remain constant, increase linearly, decrease… In such cases, we can simulate each shape of the different variants competing for dominance, as before, but excluding those considered as residuals. Now, consider *N*′ variants that behave as a peak or wave (those competing to dominate the national viral landscape) and M′ variants that can be considered as residual. The total number of variants is now *N*′ + *M*′ + 1 (to account for the one that would decline). Then, if variant *k* is one of the residual variants in the substitution, i.e., *k* = 1, …, *M*′, the dynamics of the system can be written as a function of the emerging variants:

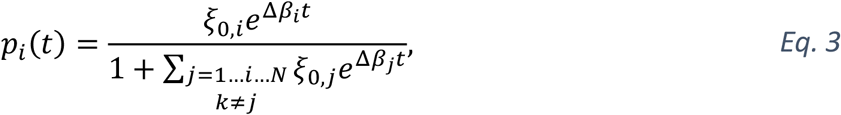

the residual variants:

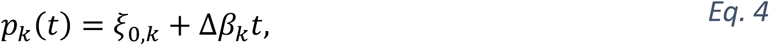

and the descendent variant:

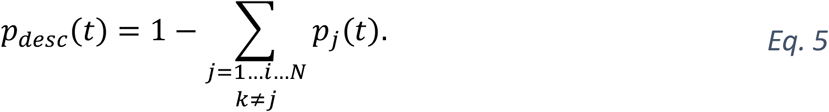

Where ξ_0,*i*_ and Δ*β*_*i*_ represent the initial ratio and the increase in transmissibility between the variant i and all other competing variants, respectively, *i* = 1, …, *N*′ and *k* = 1, …, *M*′.

These mathematical equations are employed in our computational approach to model the dynamics of SARS-CoV-2 variants, adapting the model to each substitution and country. The algorithm collects and categorizes weekly samples of each variant, defines a time window for variant substitution, and automatically determines the number of variants involved. It then identifies the residual variants and optimizes the fit for those that exhibit substitution behavior. This iterative process continues until a final substitution model is established, providing detailed statistics such as daily percentages, confidence intervals, and key metrics related to the substitution process, such as Δ*β*. Furthermore, as it extracts the estimated daily percentages of each variant in each substitution process, we can also estimate the daily cases of COVID-19 associated with a particular variant.

In the Supplementary Material we use a variety of mathematical approaches to test its robustness. Besides the aforementioned nonlinear regression, we use a Monte Carlo simulation algorithm and a weighted nonlinear regression. Both are explained in more detail in the **Suppl. Mat. Text S4**, and results can be found in the **Suppl. Mat. Table S8**.

### 2.4 Effective reproduction number analysis

We employ an empirical method for estimating the time-dependent effective reproduction number (*R*_*t*_), a critical parameter for quantifying disease transmissibility over time. *R*_*t*_ indicates the average number of people that a single infected individual will transmit the disease to at a specific point in time. It provides insights into whether an epidemic is growing or declining. If *R*_*t*_ > 1, each existing infection is expected to generate more than one new infection, implying that the spread is increasing. Conversely, when *R*_*t*_ < 1, each existing infection causes less than one new infection, decreasing the spread. We used the relationship between *R*_*t*_ and variant substitution processes to examine how the emergence of new variants affects the total number of cases. We use the definition of a previous empirical model [20], but with certain modifications [21] to account for irregularities in daily reported cases due to weekends or holidays.

Clear daily patterns led us to assign a global weight, *W*_*j,d*_, to each weekday (*d*), depending on the specific country (*j*):

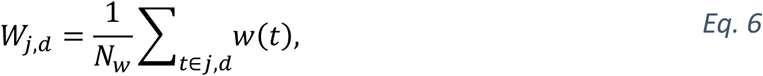

where *N*_*w*_ is the number of weeks in the series, and *w*(*t*) is the ratio of the day. This pattern takes into account weekends and, for example, December 25th, or January 1st.

Using these weights, we generate a corrected series of new cases for each country. Let’s denote *n*_*j*_(*t*) as the number of new cases in a country *j* for a given day t. We then construct the corrected series of new cases *C*_*j*_(*t*) as follows:

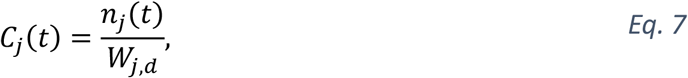

where *d* is the weekday associated with day *t*. Finally, with these new values, we can estimate the *R*_*t*_ as the ratio:

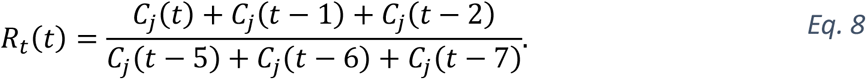

## 3. Results

In this section, we present the results using the primarily selected database (GISAID) and the nonlinear method explained in Sections 2.3, 2.4 (further information in **Suppl. Mat. Text S3 and S4**). The results of the other mathematical approaches mentioned in the previous section can be found in **Suppl. Mat. Table S8**, achieving similar results and conclusions.

### 3.1 Temporal evolution of SARS-CoV-2 variants and effective reproduction number

Here, for the temporal evolution of SARS-CoV-2 variants in Europe, we use Spain as a case study. The same analyses for other countries are presented in the Supplementary Material. The study yields three types of results: (*i*) the evolution of the VOC substitutions Alpha, Delta, and Omicron in 19 European countries; (*ii*) the evolution of the six major substitutions (Alpha, Delta, and the Omicron variants BA.1, BA.2, BA.5, and BQ.1) in 18 European countries6; and (*iii*) the dynamics of the dominant variants in each substitution process in these 18 countries. We show the first and the second results in the main manuscript and relegate the third to **Suppl. Mat. Text S7**.

The weekly evolution of the samples of each variant, the sampling percentage and the fitted substitution model for each transition period, the estimated number of reported COVID-19 cases corresponding to each variant from our model, and the evolution of the empirical effective reproduction number are shown in Figure 1 (substitution dynamics in Spain from fall 2020 to spring 2022) and Figure 2 (from fall 2020 to early 2023, also including Omicron subvariants).

**Figure 1.**
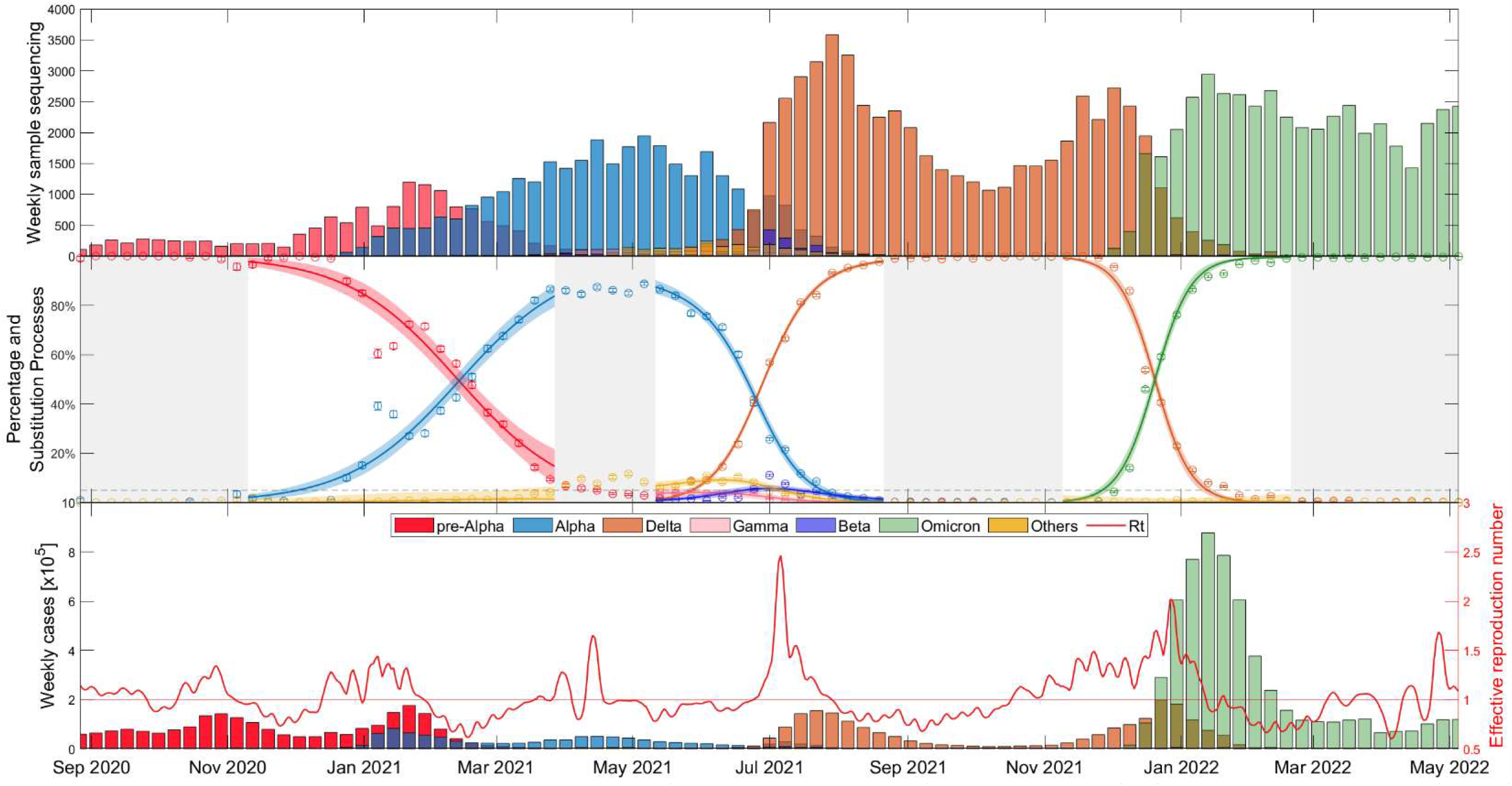
Evolutionary dynamics of SARS-CoV-2 main VOC substitutions (Alpha, Delta, and Omicron) in Spain over time: (top) Weekly sample sequencing, (middle) measured percentage data and mathematical substitution model, and (bottom) estimated weekly COVID-19 cases of each variant and the associated effective reproduction number. The 18 remaining countries and Europe figures are displayed in **Suppl. Mat. Text S5**.

**Figure 2.**
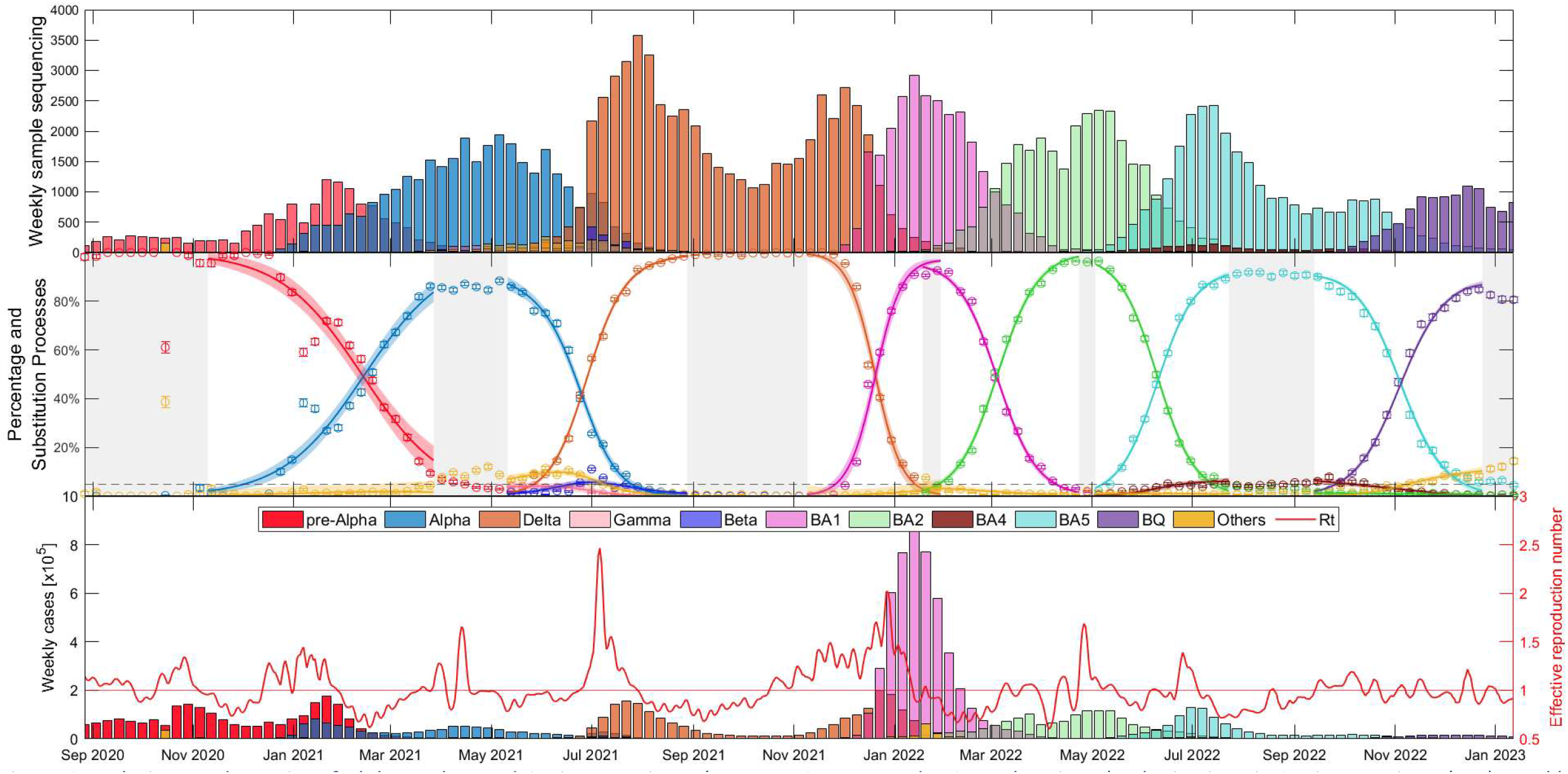
Evolutionary dynamics of Alpha, Delta, and Omicron variants (BA.1, BA.2, BA.5, and, BQ.1 subvariants) substitutions in Spain over time: (top) Weekly sample sequencing, (middle) measured percentage data and mathematical substitution model, and (bottom) estimated weekly COVID-19 cases of each variant and the associated effective reproduction number. The 17 remaining countries and Europe are displayed in **Suppl. Mat. Text S6**.

As shown in Figure 1, the time window includes the end of the dominance of the *pre-Alpha* variants (red), the slow rise of the Alpha VOC (blue), the increase in dominance of the Delta VOC (orange) along with a notable increase in cases and the *R*_*t*_ during the early summer of 2021, and after several months of dominance of the Delta variant, the rapid emergence of Omicron (green) in just over a month, during November-December 2021, along with the highest peak in case numbers.

Figure 2 shows essentially the same as Figure 1, but this time with a wider time window (fall 2020 – early 2023) and separating the Omicron variant into its different major subvariants: BA.1 (pink), BA.2 (light green), BA.4 (brown), BA.5 (light blue), and BQ.1 (purple). It is interesting to note that the duration of dominance varies significantly among SARS-CoV-2 variants and subvariants. While Alpha and Delta dominated for more than 5 and 6 months respectively, the early Omicron subvariants BA.1 and BA.2 showed relatively shorter durations of dominance, lasting only 2.5 and 3.5 months, respectively. BA.5 dominated for almost 5 months and then BQ.1 also displayed a duration of approximately 3.5 months. Additional details on the specific competition between dominant variants in each substitution process can be found in **Suppl. Mat. Text S7**, confirming observations similar to those in Figure 2.

Figures 1 and 2 provide a comprehensive view of the dynamics and evolution of COVID-19 variant substitutions in Spain, highlighting the effectiveness of the substitution model in capturing the changes in the dominant variants over time. Each of these substitution processes and figures are described in detail in **Suppl. Mat. Text S8**. The results from Spain can be used as a reference for understanding the patterns and trends in the other European countries, which are presented in **Suppl. Mat. Texts S5 and S6**.

### 3.2 Statistical analyses of transmissibility and epidemiological indicators

This section focuses on statistical evaluations of variant transmissibility of the Alpha, Delta, and Omicron, and other epidemiological indicators across the 19 European countries studied. Detailed examinations of the subsequent Omicron subvariants and two-variant analyses are relegated to **Suppl. Mat. Text S10** and **Suppl. Mat. Text S12**, respectively

#### 3.2.1 Comparing the transmissibility of the Alpha, Delta, and Omicron variants

We first examine the increase in transmissibility Δ*β* between the three main VOCs. It is important to note that other variants, although residual, may play a role during the substitution process influencing Δ*β*. For an exclusive dominant variant result consult **Suppl. Mat. Text S12**, but these variants rarely exceed 10% of the national landscape.

Figure 3 shows Δ*β* results for the 19 European countries and Europe as a whole (sum of variants for each week and country), as well as a boxplot of the fitted Δ*β* for all 19 countries. There is a consistent trend across countries: ∆*β*_Alph_ < ∆*β*_*Delta*_ < ∆*β*_*Omicron*_. This suggests that the Omicron variant spread more rapidly than both the Alpha and Delta variants.

**Figure 3.**
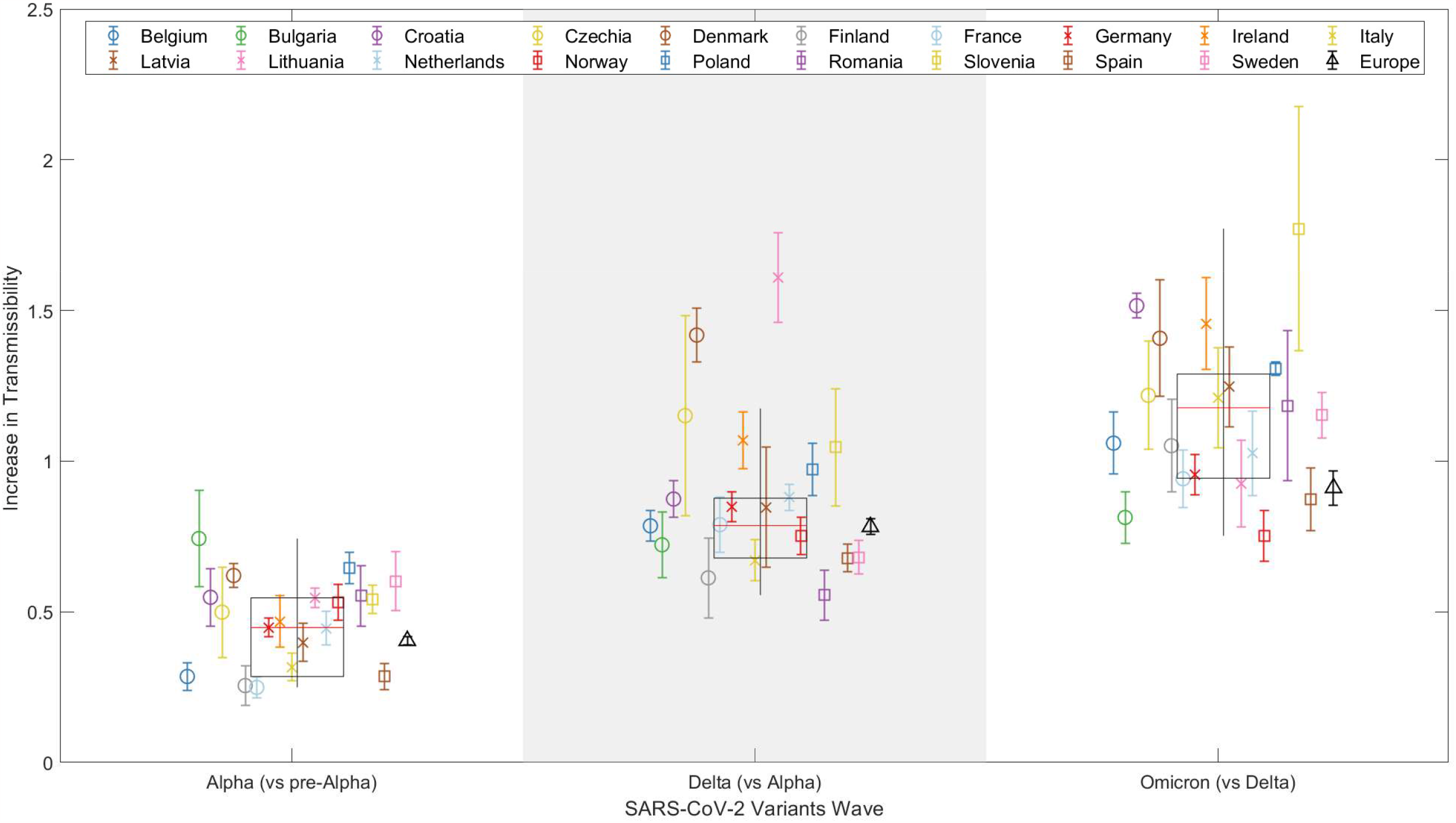
Representation of the increase in transmissibility (∆*β*) for 19 European countries and Europe (see legend) as a combination of the studied countries, based on the results for the Alpha, Delta, and Omicron VOCs. Weighted boxplots (black boxes) are also included to provide statistical summary. The x-axis distinguishes the three SARS-CoV-2 VOC substitutions, while the y-axis displays the Δ*β* value for each substitution and country.

Figure 3 also shows weighted boxplots (black boxes) that account for the error associated with the Δ*β* of our nonlinear regression models. Each Δ*β* value has an associated error, and we use the inverse of that error as the weight. By assigning higher weights to Δ*β* parameters with smaller errors, we ensure that the parameters that we estimate with greater precision have a greater impact on our summary statistics. This is especially valuable when the data distribution might be skewed or outliers could disproportionately affect the results. The calculations were done without Europe as a whole, but with the 19 countries used in the analysis. Each box is centered on the median of the data (red line), with the edges of the box indicating the first and third quartiles. The whiskers extend vertically from the box to demonstrate the range of the data, indicating variability outside the upper and lower quartiles, thus providing a sense of the spread and skewness of the data. Points outside the vertical lines, can be considered outliers, thus only Denmark and Lithuania can be considered outliers in the Alpha-Delta substitution. This does not mean that both substitutions are wrong, but that in these two countries the emergence of the Delta variant did not follow the global trend of the others.

Note that the data point for Europe (represented by the black triangle in Figure 3) sits below the median (red line of the boxplot) for all substitutions, although they are nearly equal during the Alpha to Delta substitution. This is attributed to how the point for Europe is calculated, integrating all sequenced samples during these periods, and thus strongly affected by asynchrony among substitutions, as we discuss below in Section 3.3. On the contrary, the boxplot accounts for the distribution of the individual fittings and this is not affected by the date of the substitution onset. Table 1 presents these results.

#### 3.2.2 Relationship between Δ*β* and initial day of variant emergence

Variability in Δ*β* exists both between variants and between countries for the same variant. Factors like variant entry timing, country characteristics, vaccination rates, and current epidemiological conditions can influence these variations.

Figure 4 shows the Δ*β* values relative to the entry date of the dominant variant for each of the three waves of new VOCs. The entry date is defined as the day when the percentage of the new variant exceeded 5%, according to the fitted model. Here, a trend emerges: higher Δ*β* values are associated with later entry dates for the new variant in a given country. This could be explained by a higher probability of multiple simultaneous entries of the new variant as its prevalence increases in other countries. However, the trend is not perfectly linear, which is to be expected given the various characteristics of the different countries.

**Figure 4.**
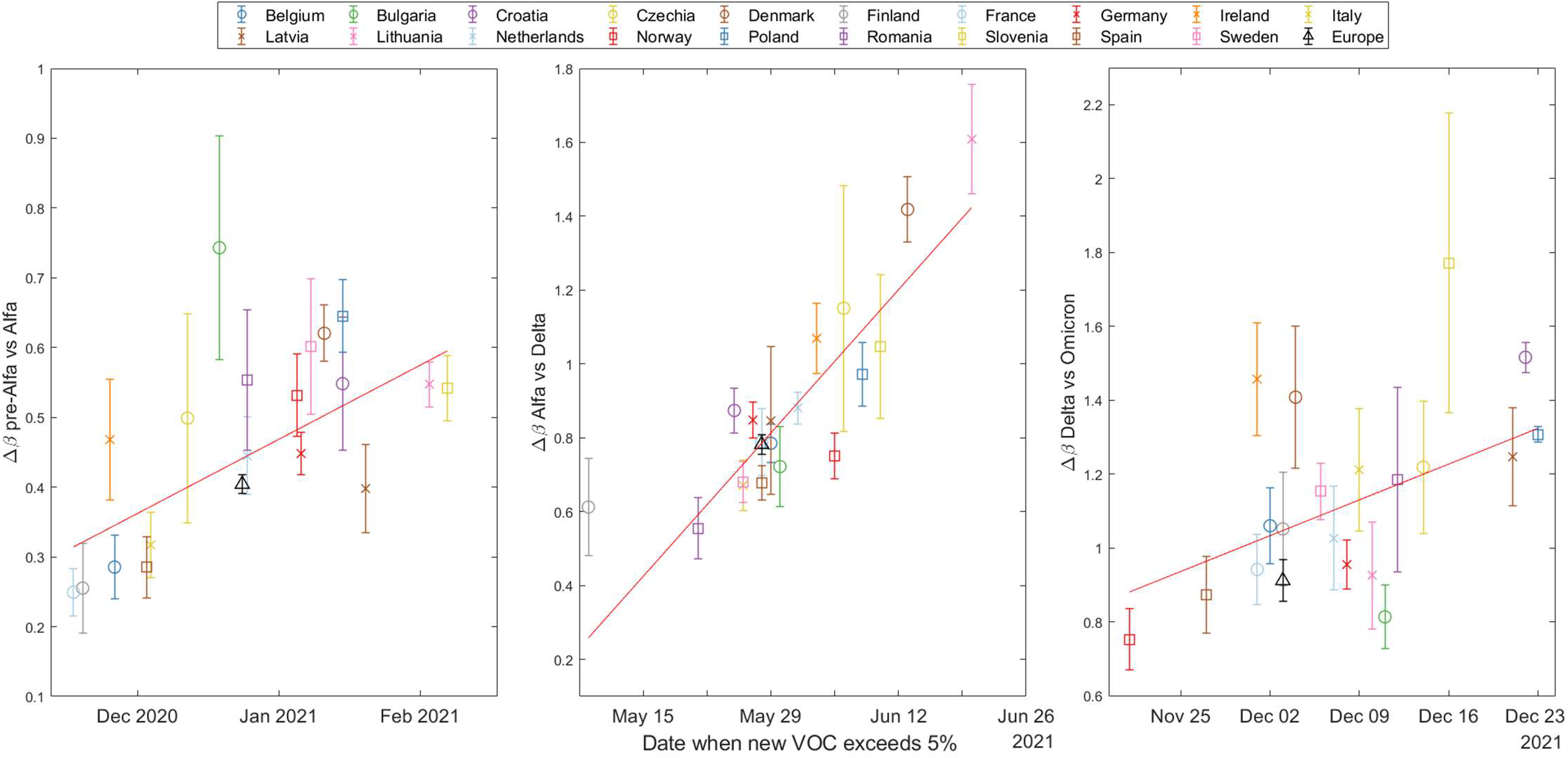
The increase in transmissibility (Δ*β*) based on the date the emerging variant (Alpha on top, Delta in the middle, and Omicron at the bottom) exceeded 5% according to our substitution model. Note that the y-axis are different and then the slopes can not be compared between plots.

To quantify these trends, we performed Spearman tests, and calculated the coefficient of determination *R*^2^; see **Suppl. Mat. Text S10** for a more detailed explanation of these statistical tests. The results are shown in Table 2.

**Table 2.**
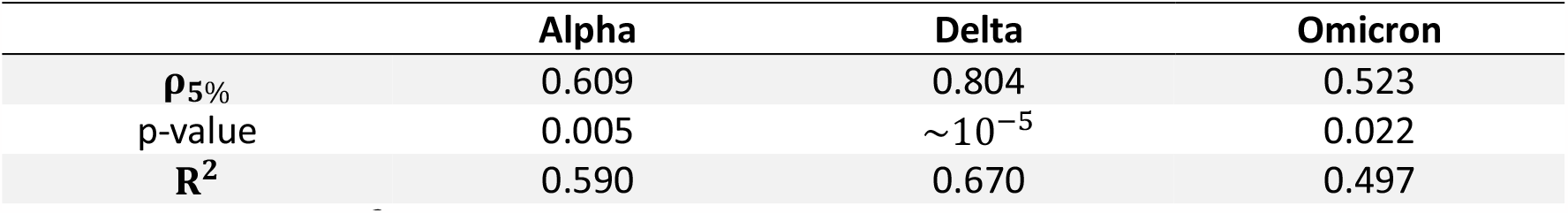
Statistical results for trend analysis across three variants transitions: Alpha, Delta, and Omicron.

|  | Alpha | Delta | Omicron |
| --- | --- | --- | --- |
| $\rho_{5\%}$ | 0.609 | 0.804 | 0.523 |
| p-value | 0.005 | $\sim 10^{-5}$ | 0.022 |
| $R^2$ | 0.590 | 0.670 | 0.497 |

As is to be expected, *R*^2^ is not very large, reflecting the complex interplay of real country-specific factors, but the fact that all three p-values are below 0.05 indicates that the variables (Δ*β* and entry date) are correlated, although not linearly.

#### 3.2.3 Influence of country surface area on Δ*β*

We divide the countries of the study into two distinct clusters, in order to better address the role of the surface area: smaller and larger countries. These clusters are categorized based on a geographic area threshold of 200,000 km^2^. This criterion separates larger countries, referred to as cluster 2, including Romania, Italy, Poland, Finland, Germany, Norway, Sweden, Spain, and France (listed in increasing order of size), from the smaller countries (cluster 1) including Bulgaria, Czech Republic, Ireland, Lithuania, Latvia, Croatia, Denmark, Netherlands, Belgium, and Slovenia (listed in decreasing order of size).

This analysis does not provide a clear statistical trend for any substitution or across countries. However, by separating the European countries into two clusters, a discernible pattern emerges as shown in Figure 5. For each VOC transition—Alpha, Delta, and Omicron— the median Δ*β* parameter is consistently lower for countries with a larger geographic area (cluster 2). This suggests that factors such as population (**Suppl. Mat. Text S9**) and geographic area may influence the rate of global variant substitution. Specifically, in larger countries, an asynchronous emergence of the same variant across different regions could potentially affect the substitution process at the national level, leading to an apparent slower increase in transmissibility. This may also explain the consistently lower Δ*β* value for Europe compared to the median of individual countries, as discussed in Section 3.2.1. Further insights into this issue are explored in Section 3.3.

**Figure 5.**
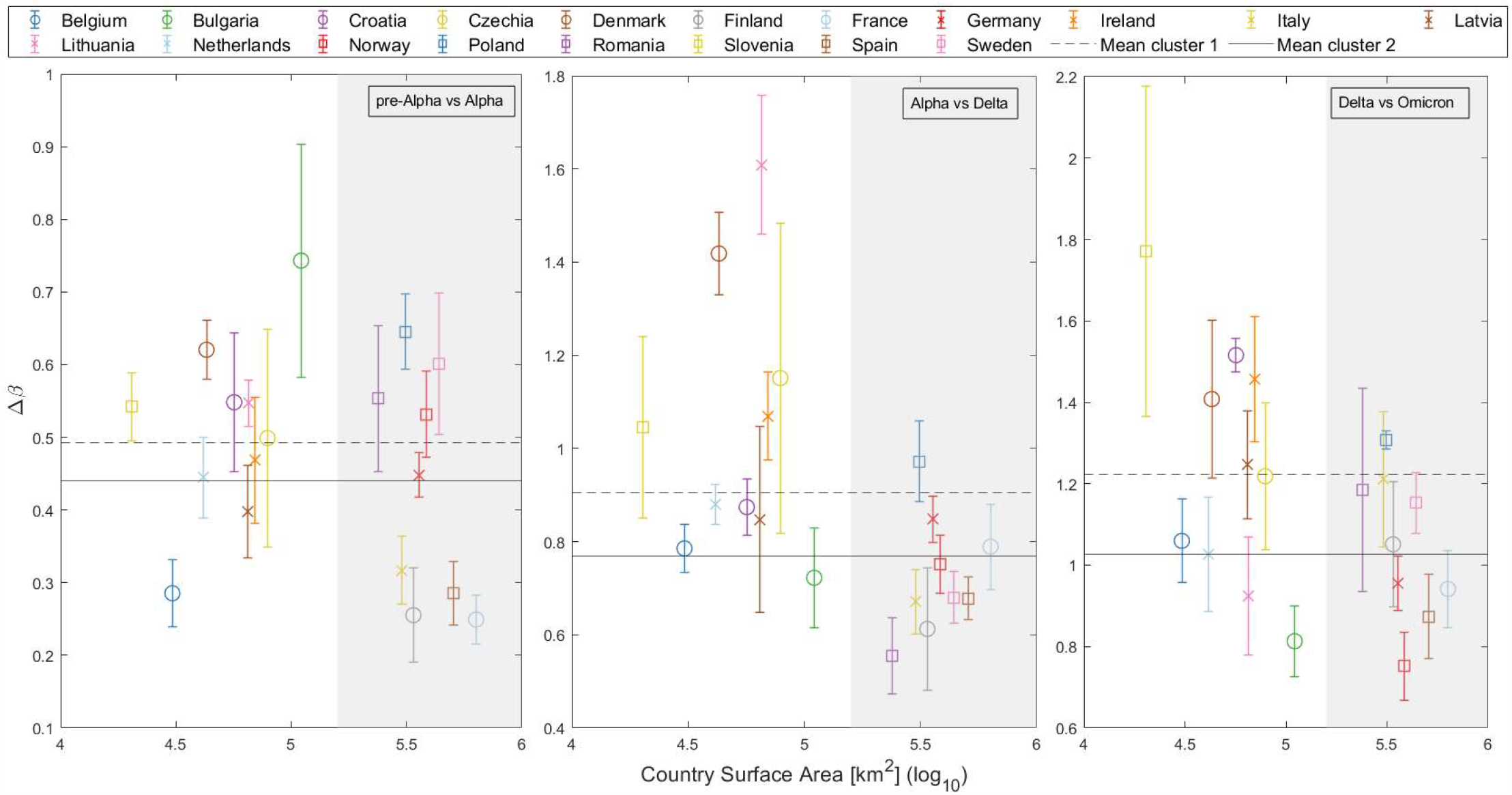
Increase of transmissibility (Δ*β*) plotted against the log of the country’s surface area, distinguishing two primary clusters: smaller countries (cluster 1) on a white background, and larger countries (cluster 2) on a gray background. Mean Δ*β* for both clusters are depicted in each substitution with horizontal lines (cluster 1: Dashed line; cluster 2: Solid line).

#### 3.2.4 Δ*β* in relation to the percentage of fully vaccinated population

Vaccination is a key parameter in understanding the global dynamics of SARS-CoV-2. Vaccines should reduce disease severity, transmission rates, and variant susceptibility. However, the relationship between vaccine coverage and the transmissibility of new variants probably depends on several factors, making the relationship complex.

At the time of the Alpha variant emergence (December 2020 – January 2021), vaccination programs were not yet widely implemented in Europe. For the Delta variant, our analysis presents an interesting trend. As Figure 6 illustrates, countries with higher vaccination rates showed a greater Δ*β* of the Delta variant. This result would suggest a higher spread advantage of the Delta variant over Alpha among vaccinated people, which could be explained by a higher vaccine escape of the former, although other factors like natural immunity and non-pharmacological interventions could play an important role. In this analysis, consistent with our previous studies on this substitution, Denmark and Lithuania were excluded due to their outlier status. Statistical testing revealed a Spearman correlation coefficient of 0.5491 with a p-value of 0.0164, indicating a moderate positive correlation between the vaccination coverage and the increase in transmissibility of the Delta variant.

**Figure 6.**
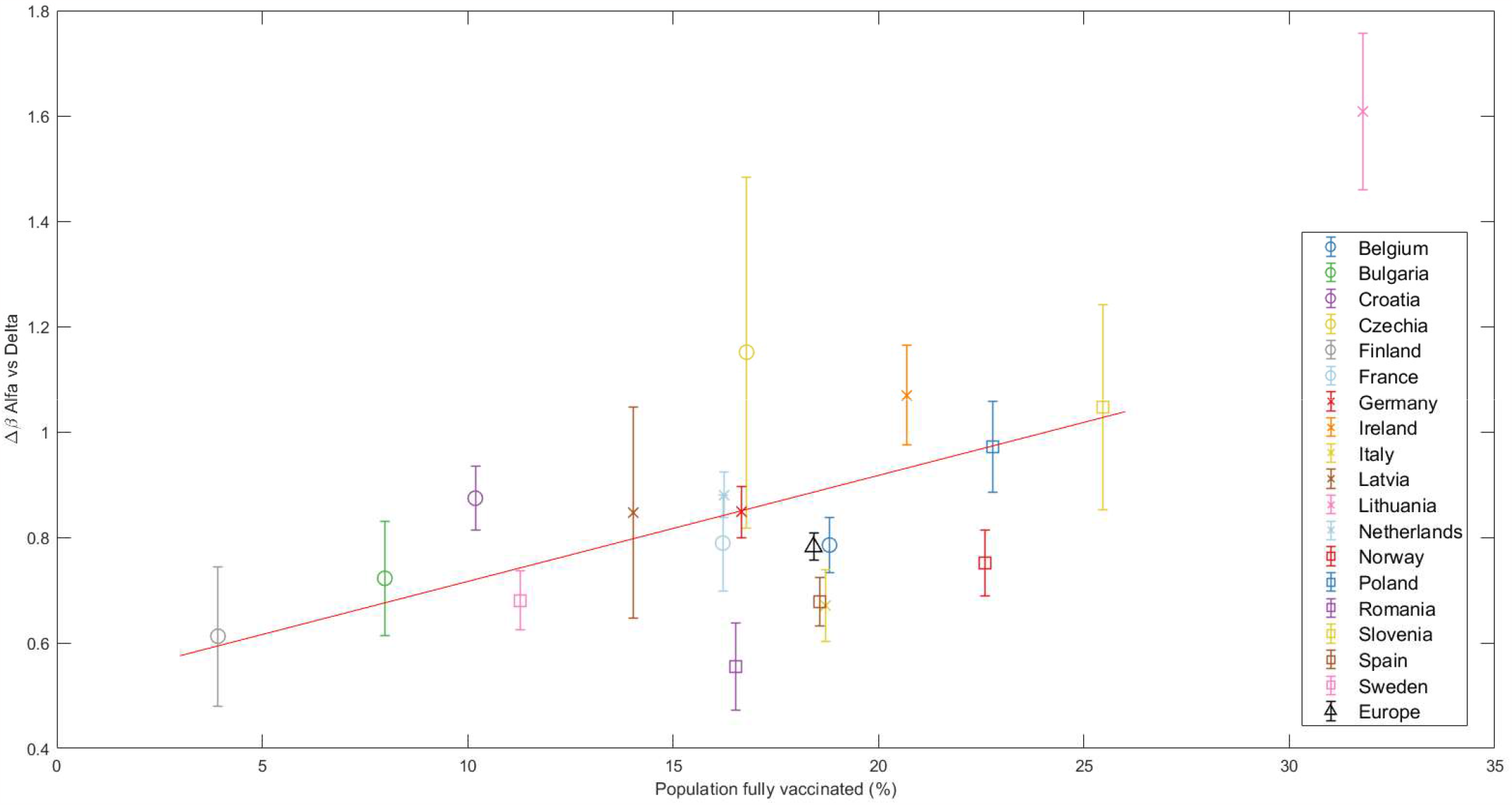
The increase in transmissibility Δ*β* is plotted against the percentage of fully vaccinated individuals at the beginning of the Alpha-Delta substitution (Delta > 5%). The red line marks the linear regression, clearly indicating an increasing trend confirmed by the Spearman test. Other substitutions are presented in the Supplementary Material Text S9.

Finally, for the Omicron variant, no clear trend was observed with respect to the vaccination coverage, which is an expected result given the higher immunity level of the population. These results and those for the Alpha substitution can be found in **Suppl. Mat. Text S9**.

#### 3.2.5 Trends in the effective reproduction number during variant substitutions Finally, a key parameter in understanding the day-to-day progression of a pandemic is the effective reproduction number, or *R*_*t*_

Figure 7 shows the relationship between the effective reproduction number at the beginning of the substitution (when the old variant dominates, x-axis) and the end (when the new variant dominates, y-axis) for each of the three VOCs substitutions. The data points were calculated by taking the days when the new variant was at a specific percentage (based on our daily substitution model) and calculating the average. These percentages were set at 20-40% for the start and 60-80% for the end of the substitution. We did not choose values lower than 20% or higher than 80% because in most countries, the Alpha variant never exceeded 90% of the total sampling. Thus, the ranges were symmetrical and consistent for each of the three substitutions. Note the left side of Figure 7 (*pre-Alpha* vs Alpha substitution) is missing some data points because the substitution does not reach 80%. The diagonal dashed line in the figure represents the 1/1 boundary. We observe that most of the points fall above this line, i.e., suggesting that *R*_*t*_ generally increases with the entry of a new variant.

**Figure 7.**
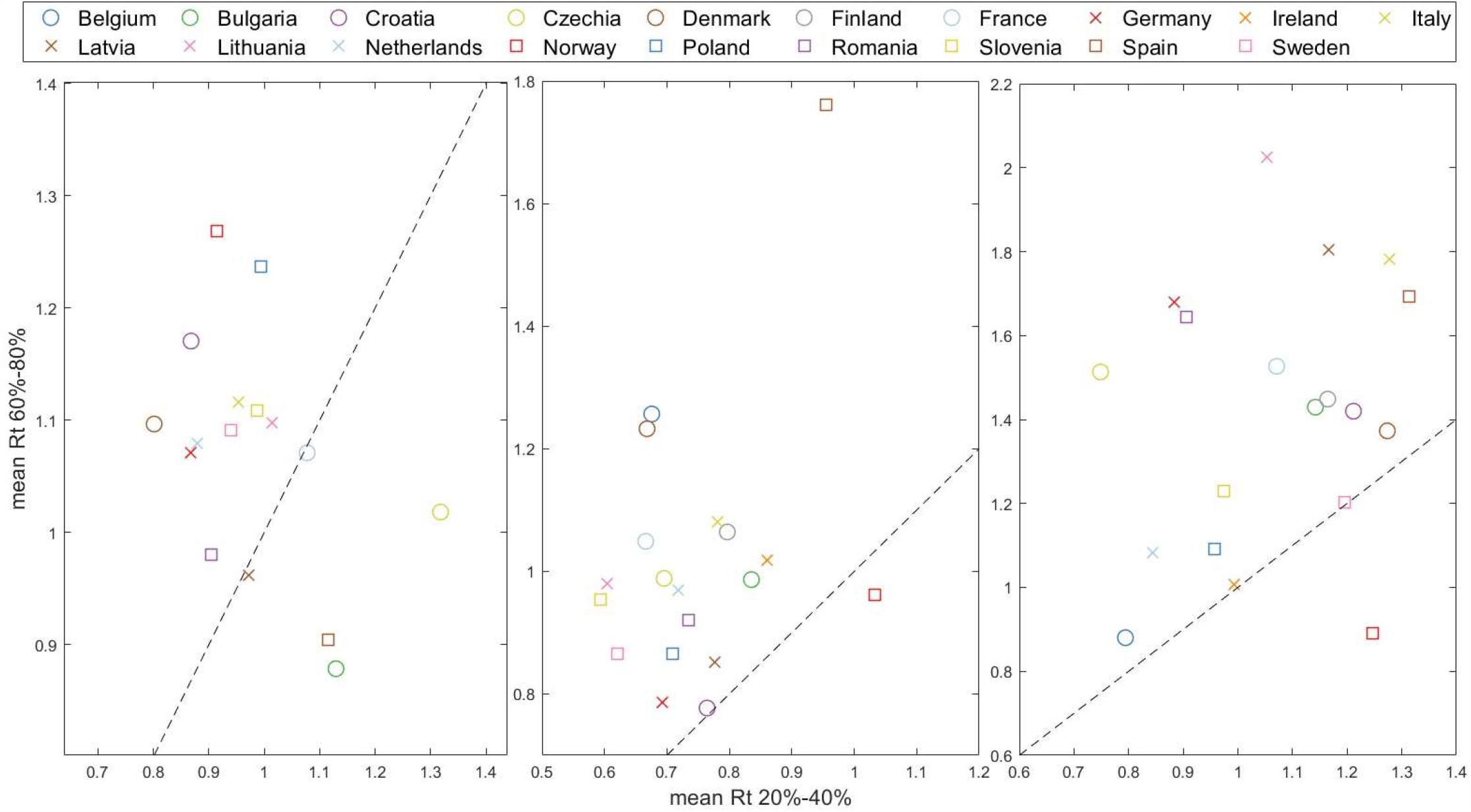
Effective reproduction number mean 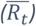 at the end of substitution (accounting for 60%-80% of the emerging variant) versus 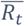 at the beginning of the substitution (20%-40% of total). The dashed line indicates the 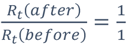 threshold, where points above signify an acceleration of the pandemic spreading at the end of substitution.

### 3.3 The impact of different outbreaks within a single country: a case study of Spain

In the previous section, we observed that larger countries (Figure 5) generally have slower rates of increase in transmissibility Δ*β* and when we aggregate all sequenced samples and derive results for Europe (Figure 3), they also tend to fall below the median. In this section, we aim to explore how regional outbreaks within a large country like Spain can impact its overall viral landscape.

Spain is divided into 17 regions known as Autonomous Communities (*Comunidades Autónomas* or CCAA in Spanish) and 2 autonomous cities. We used data obtained from the GISAID website, which provides comprehensive information about each sample: day, lineage, location, age… We focused on the Alpha vs Delta substitution, as there were no major restrictions in the country, the number of weekly samples was high enough to apply the substitution model to individual CCAA, and the substitution was slow enough to observe the expected effect.

We studied those CCAA that exceeded 1000 sequences during the months of May and June and merged some medium-sized neighboring CCAA into a single region to reach this threshold. These CCAA are Andalusia (south), Balearic Islands (Mediterranean Sea), Castile and León (north central), Catalonia (northeast), Valencian Community + Murcia (along the Mediterranean coast on the east side), Madrid (center), and Navarre + Basque Country (north of Spain). The daily percentage results calculated with the mathematical model from Section 2.3 for these 7 groups are shown in different colors in Figure 8 (left). This figure shows how regions in the northeast and east of Spain were the first to show an increase in the Delta variant. It gradually spread to other parts of the country, resulting in a time gap of more than two weeks between the emergence of the same variant. Looking at the shape of the curve for the whole country, as indicated by the dashed black line, we can predict that the calculated increase in transmissibility for Spain will be lower than the CCAA average. The percentage curve of Spain begins to rise with Catalonia and the Valencian Community, but as the weeks go by, the curve of Spain relaxes and approaches its maximum at the same time as the last CCAA do. In summary, in large countries, the curve slows down from its initial increase driven by the first regions experiencing variant substitution as these regions reach complete substitution while the rest lag behind.

**Figure 8.**
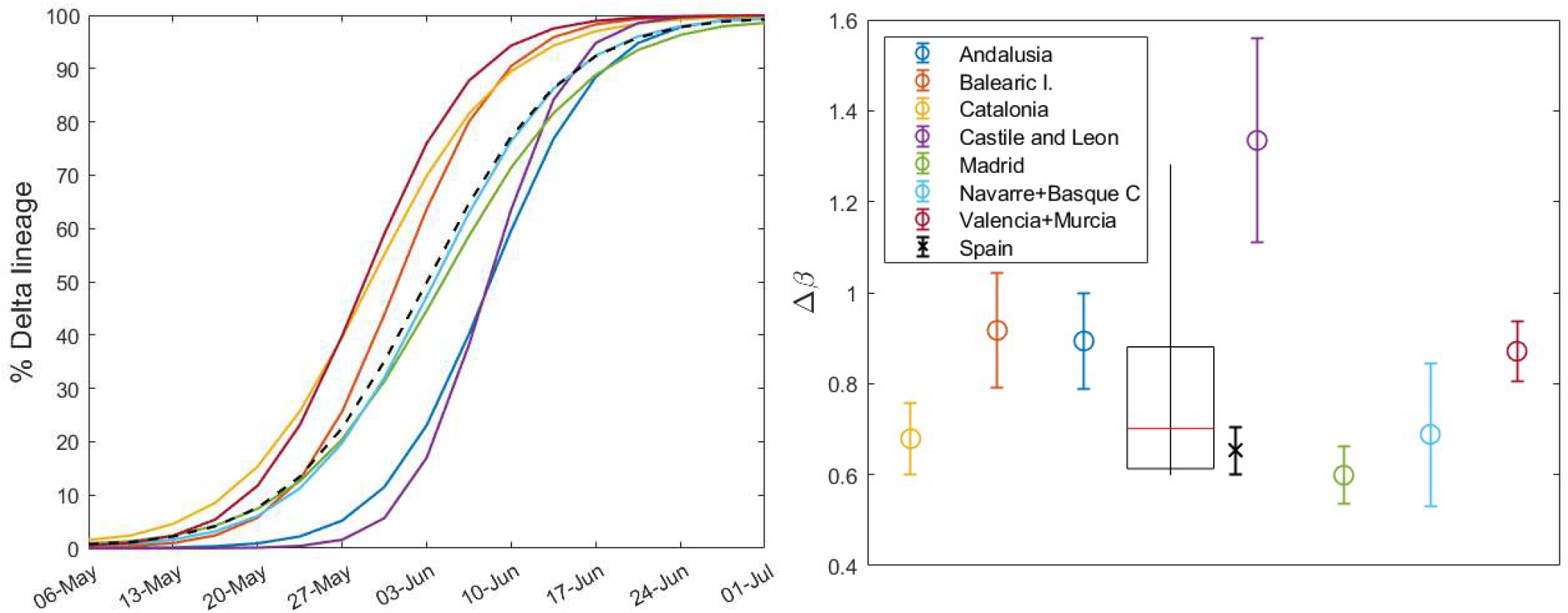
(Left) Daily percentage (without error margins for clarity) of the Delta variant during the substitution process, calculated for different CCAA with n_sample_ ≥ 1,000 during these months. The dashed curve corresponds to the country, calculated solely from the weekly sample sums from the seven regions shown here. (Right) Increase in transmissibility calculated by the substitution model for each region (circles) and for Spain (cross), with a boxplot showing the median, first and third quartiles, and outliers.

The results of Δ*β* from our model are shown in Figure 8 (right): the circles stand for CCAA and the cross represents Spain. Note that the result for Castile and León exceeds the interquartile range and will be considered an outlier. The weighted boxplot was calculated in the same way as in Section 3.1. As previously suggested, the result for Spain (black cross), as the sum of the sequenced samples taken weekly in each region, is lower than what we would obtain from the median of the different regions (red line in the boxplot). This supports the previous statements and reinforces the findings of the preceding sections. Note that the increase in transmissibility for Spain, Δ*β*_*Spain*_ = 0.65 ± 0.05, does not correspond with that of the previous Section 3.2, Δ*β*_Spain,3.2_ = 0.68 ± 0.04, since we have not taken into account all the regions or CCAA.

### 3.4 Mathematical model with two Omicron subvariants strongly competing and plausible short to medium term scenarios

After the emergence and dominance of Omicron in Europe, the SARS-CoV-2 viral landscape has been characterized by frequent competition, typically involving more than two subvariants. But can we use our substitution model to determine which of the competing variants will ultimately dominate the viral landscape of a country? Our mathematical and computational model returns the parameters Δ*β* and *ξ*_0_ based on initial and final conditions set for consistency across countries and substitutions (see **Suppl. Mat. Text S3**). For Δ*β*, which may give us an idea of a short to medium subvariant dominance scenario, the time window is important.

Let us focus only on the last substitution of this study period: during the last quarter of 2022, Europe experienced a shift from the BA.5 variant to BQ.1 and, rapidly, to XBB subvariants (especially XBB.1.5). The Δ*β* values from our model (Figure 2), which indicate the increase in transmissibility, showed that BQ.1 and XBB are closely competing for dominance, see Table 3 (left columns). We see no other substitution or country with such closely matched figures (see **Suppl. Mat. Table S9** to compare Δ*β* values for all countries and substitutions).

**Table 3.** Parameters for BQ.1 and XBB during substitution, taking into account their individual peak growth periods.

| Country | $\Delta\beta \pm \varepsilon_{\Delta\beta}$ | | | |
| --- | --- | --- | --- | --- |
|  | BA.5 vs BQ.1 substitution |  | BA.5 vs XBB substitution<br>(extended weeks) |  |
|  | BQ.1 | Others (incl. XBB) | BQ.1 | XBB |
| **Belgium** | 0.393±0.023 | 0.368±0.048 | 0.352±0.032 | 0.827±0.046 |
| **Croatia** | 0.257±0.025 | 0.285±0.038 | 0.241±0.032 | 0.790±0.083 |
| **Czechia** | 0.223±0.048 | 0.318±0.054 | 0.238±0.048 | 0.693±0.068 |
| **Denmark** | 0.364±0.027 | 0.421±0.040 | 0.281±0.035 | 0.661±0.044 |
| **Finland** | 0.281±0.026 | 0.222±0.052 | 0.231±0.037 | 0.663±0.052 |
| **France** | 0.348±0.015 | 0.348±0.050 | 0.322±0.018 | 0.807±0.027 |
| **Germany** | 0.223±0.016 | 0.303±0.037 | 0.212±0.014 | 0.594±0.022 |
| **Ireland** | 0.371±0.038 | 0.413±0.061 | 0.381±0.044 | 0.850±0.050 |
| **Italy** | 0.283±0.022 | 0.288±0.057 | 0.260±0.023 | 0.682±0.040 |
| **Netherlands** | 0.312±0.018 | 0.327±0.030 | 0.307±0.027 | 0.700±0.033 |
| **Norway** | 0.311 ±0.048 | 0.279±0.077 | 0.271±0.062 | 1.02±0.207 |
| **Poland** | 0.400±0.059 | 0.351±0.076 | 0.389±0.066 | 0.778±0.075 |
| **Romania** | 0.418±0.129 | 0.463±0.158 | 0.355±0.098 | 0.725±0.118 |
| **Slovenia** | 0.340±0.035 | 0.492±0.059 | 0.328±0.048 | 0.672±0.059 |
| **Spain** | 0.514±0.026 | 0.554±0.055 | 0.484±0.038 | 0.933±0.044 |
| **Sweden** | 0.352±0.023 | 0.454±0.045 | 0.285±0.037 | 0.662±0.054 |

We repeated our analysis specifically for this substitution. Here, instead of grouping XBB within the “Others” package as in previous sections, we separated it into an independent group (samples sequenced and classified as both generic XBB and XBB.1.5). We also did the same with the BA.2-like (including BA.2 and BA.2.75) because it was strongly competitive. This new analysis was conducted over a larger time window to observe changes in ∆*β*: initially, we assumed that the final day of the simulation coincides roughly with the peak of the BQ.1, and now we assume that the final day of the time window corresponds to the peak of XBB. In this analysis, Bulgaria, Latvia, and Lithuania were excluded due to discontinuities in their reporting of sequenced samples. The results are shown in Table 3 (right columns). Here we can see how the parameter Δ*β*_BQ.1_ remains relatively stable with an average variation between the columns of about 9%. On the other hand, the Δ*β*_XBB_ parameter varies considerably, always increasing in the last column. The first double column (substitution of BA.5 for BQ.1) is somewhat truncated and doesn’t adequately represent XBB; nevertheless, it shows us that the BQ.1 variant has a clear competitor. By setting the correct time windows, we obtain the real results for all variants in this last complex substitution.

Figure 9 illustrates this for two extreme cases, Czechia, where it is immediately apparent that the “Others” category (including XBB) will quickly dominate (Δ*β*_*BQ*.1_ = 0.223 ± 0.048 < Δ*β*_*Other*_ = 0.318 ± 0.054), and Poland, where the same conclusion may not be initially obvious (Δ*β*_*BQ*.1_ = 0.400 ± 0.059 > Δ*β*_*Othe*_ = 0.351 ± 0.076), but eventually occurs. This is significant because by comparing the real-time increase in transmissibility, we can understand the current state of the viral landscape of a country. In fact, we could predict the prevalence of currently circulating variants in the short to medium term. For a comprehensive overview of the XBB patterns, equivalent figures for other European countries are provided in Supplementary Material Text S13.

**Figure 9.**
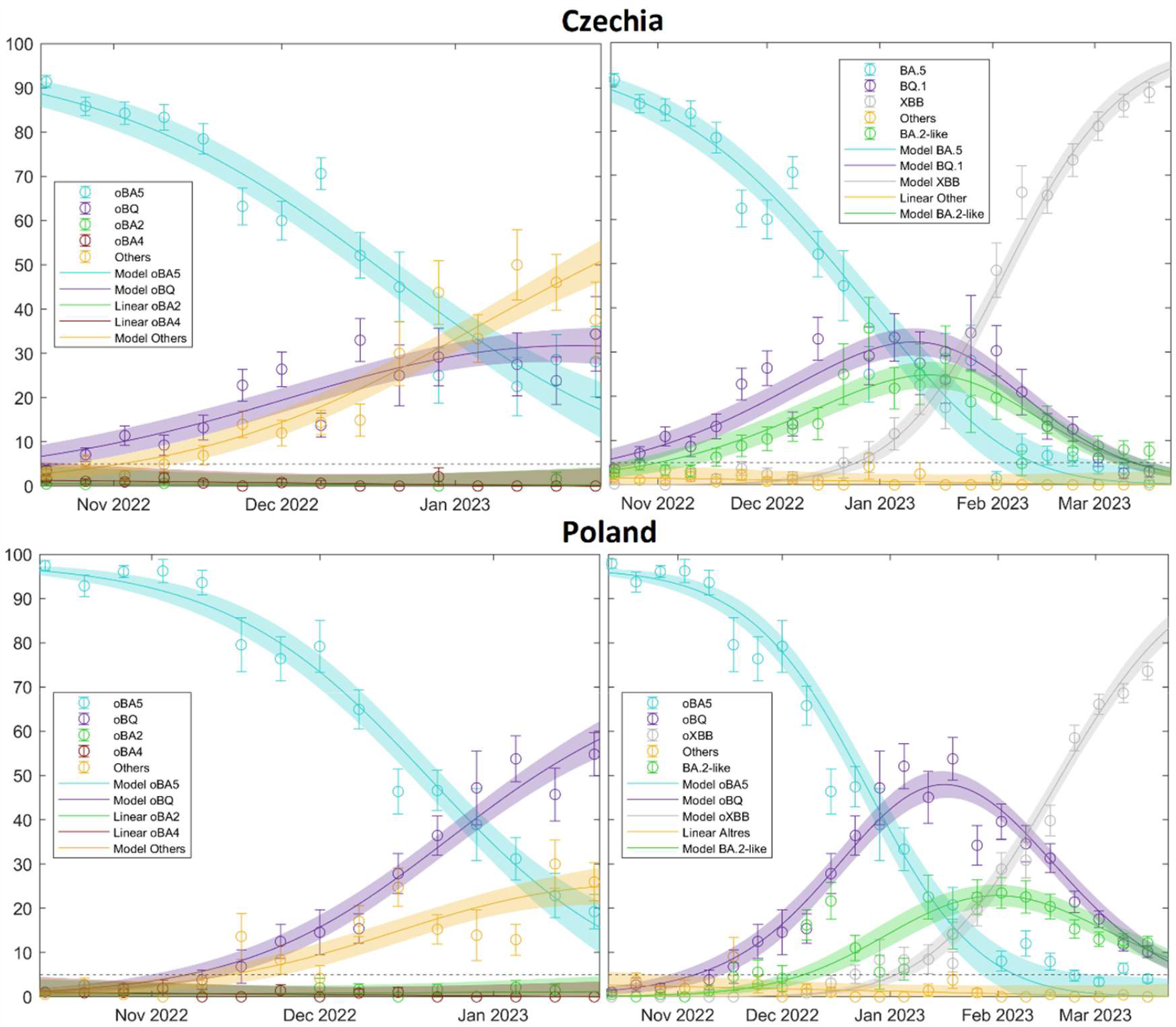
Comparison between Czechia (top) and Poland (bottom) during two-time windows: September-January (left) and September-March (right) for a substitution event marked by strong competition between variants. The Omicron BA.5 variant sees a rapid decline, being replaced primarily by BQ.1 and, in Czechia’s case, also by BA.2-like. The extended time frame reveals the ascent of the Omicron XBB subvariant towards February-March 2023.

## Discussion

Our study presents a comprehensive analysis of SARS-CoV-2 variant substitution across 19 countries using publicly available data from the GISAID database. Although the model is limited by the available data, hence focusing on a specific number of countries, the flexibility of the model allows it to adapt to different variant transition scenarios. Nonlinear regression was chosen for its speed and accuracy.

The results presented in Figures 1 and 2 for Spain (and Figures S2-S40 for the other European countries) demonstrate the robustness and adaptability of our mathematical approach in capturing the nuanced dynamics of SARS-CoV-2 variant substitutions. The model is capable of closely matching the experimental data, even in scenarios where the availability of sequenced samples is limited and multiple competing variants are spreading simultaneously. Thus, our model can provide an accurate representation of real-world variant dynamics and serve as a valuable tool to complement variant sampling in genomic surveillance.

The temporal and spatial evolution of SARS-CoV-2 is evident through our analyses of the increase in transmissibility Δ*β* for the Alpha, Delta, and Omicron variants, which show a consistent trend across European countries, with the Omicron variant exhibiting significantly higher transmissibility (Figure 3) [11], [22]. It is noteworthy that the results for the Omicron variant exhibit a higher degree of dispersion compared to those for the Alpha and Delta variants. When the Omicron variant emerged in late 2021, many European countries had relaxed containment strategies, including lockdowns and travel restrictions, and there were significant differences in vaccination coverage across countries. This heterogeneity, along with the emergence of immune escape variants and possibly other factors, may have contributed to the observed variability in the increase in transmissibility for the Omicron variant. Additionally, the pattern is more heterogeneous across the different Omicron subvariants, though each new dominant variant is more transmissible than the previous one (**Suppl. Mat. Text S10 and Figure S65**).

Generally, our data suggest that later-entrant variants spread more rapidly (Figure 4). We hypothesize that this may be mainly attributed to a larger initial number of infections, possibly due to multiple entry points, facilitated by the prolonged circulation of the variant elsewhere. This broader initial presence may accelerate transmission within the country. Interesting results also arise when examining different Omicron subvariants: while BA.1 and BA.5 seem to exhibit similar patterns, BA.2 and BQ.1 appear to be insensitive to this factor (details can be found in **Suppl. Mat. Text S10**).

Countries of different sizes display distinct substitution patterns, with larger countries systematically experiencing slower variant substitution (Figure 5). This phenomenon could be attributed to multiple epidemiological curves at regional levels separated in time. Smaller countries exhibit relatively rapid variant substitution, potentially due to the difficulty to have geographically separated outbreaks. However, this does not mean that homogeneous countries will all experience the same rate of increase in transmissibility; variations can occur due to differences in social networks. To corroborate the impact of geographical factors, we studied Spain in detail—a study that could be applicable to other large countries or even to Europe as a whole. Section 3.3 underscores the importance of understanding regional differences in the spread of SARS-CoV-2 variants. A low Δ*β* in larger countries can be attributed to the sequential peaks of outbreaks in different cities or regions. The emergence of new variants in a country begins with initial outbreaks in some areas and ends when the last regions reach their peak, resulting in a smoother and more delayed curve on a country-wide scale, which subsequently leads to lower Δ*β* values. Furthermore, we demonstrate that the country-wide Δ*β* value is smaller than the Δ*β*_median_ value obtained from individual regions within the same country (Figure 8). The same results are obtained for Europe and for individual countries. Again, interestingly, the same country surface area-pattern than before occurs within the Omicron subvariants: BA.1 and BA.5 follow the same result but BA.2 and BQ.1 are different from the rest (see **Suppl. Mat. Text S10**).

An important aspect to consider is the interplay between vaccination campaigns and the emergence of new variants [23] [24] [25] [26]. When a new variant emerges, against which prior immunization or vaccines are less effective, one could expect that it will encounter a large population of susceptible individuals, thus achieving increased transmissibility. However, this relationship is complex due to several factors, such as the immunity escape mechanisms of new variants and the non-pharmacological interventions in place at the time. In our study, we only observe a correlation between vaccination rates and the increase in transmissibility of the Delta variant (Figure 6Figure 6). However, no such correlation is evident for the Omicron subvariants (**Suppl. Mat. Text S10** and Figures S80-S83). This suggests that Omicron, a variant with high levels of immune escape (as well as its subvariants), can infect and spread widely regardless of the number of vaccinated individuals. Therefore, while vaccines may not halt the spread of the virus, they do mitigate its effects.

Interestingly, our study consistently shows a higher effective reproduction number, *R*_*t*_, toward the end of a variant substitution cycle for the majority of countries and substitutions (Figure 7). This suggests that so far new variants retain the capacity to infect in spite of preexisting collective immunity. This observation has been noted in other papers [19], [27], [28], but we have extended previous analyses by comparing the values at the beginning and at the end of the substitution process for an emerging variant. Similar results are observed for the Omicron subvariants (**Suppl. Mat. Text S10 and Figures S86-S88**) suggesting a substantial increase in cases following the emergence of a new subvariant.

Finally, our model has notably proven its reliability in scenarios involving multiple competing variants, as illustrated by our analysis of the BQ.1 and XBB Omicron subvariants, among others (Figure 9). It also underscores the importance of appropriately selecting time windows. By doing so, we can not only depict the intricate dynamics underlying variant competition, but also forecast which variant will become dominant, when it will peak, and its likely prevalence.

Despite its significant contributions, this study has some limitations. The analysis is limited to GISAID data, which restricts our geographic scope to 19 countries. While the model has performed well under various scenarios, its performance with data from other sources (to expand coverage to non-European countries) or larger sample sizes remains to be evaluated. In addition, the rapidly evolving nature of the virus requires constant updating, making long-term predictions unfeasible within the framework of our current mathematical approach. Moreover, the relationship between the transmissibility of different variants and the vaccination rates of a country is quite complex. The introduction of vaccines has led to a decrease in non-pharmacological measures, initially affecting the Alpha variant and subsequently showing a correlation with the emergence of the Delta variant based on vaccination rates. Nonetheless, for the different Omicron subvariants, the level of vaccination appears to have no discernible impact on the transmissibility of the variant, though it may influence the severity of the illness.

This study not only enriches our understanding of the evolutionary trajectories of SARS-CoV-2 variants, but also but gives an idea of the homogeneity in how the pandemic has unfolded in Europe, contrary to what other articles aiming at a global study of the planet suggest [14], giving a common response strategy for possible new waves of infection.

## 5. Conclusions

Our study offers valuable insights into the pattern of variant substitutions of SARS-CoV-2 across 19 countries. The main findings can be summarized in various points. (*i*) The continuous evolution of the virus has resulted in an increasing trend of its transmissibility from the Alpha variant to Delta and subsequently to the various Omicron variants. (*ii*) Later-emerging variants tend to spread more rapidly upon their entry into a population, what can be explained by a higher chance of multiple entries. (*iii*) The spread pattern of COVID-19 variants shows significant variations across countries of differing sizes, with a slower substitution pattern in larger countries. (iv) A significant correlation exists between vaccination rates and the speed of Delta variant propagation following the period of Alpha dominance. However, this correlation is absent with the Omicron subvariants. This highlights the complex relationship between the spread of different variants and vaccination efforts, emphasizing the urgent need to reassess vaccination strategies considering emerging new variants. This absence of correlation may also be linked to a reduction in non-pharmacological measures. (v) *R*_*t*_ values tend to increase with a new variant substitution process, emphasizing the spread potential of new variants. (vi) The model shows robustness even in scenarios of multiple competing variants, accurately representing the underlying dynamics of variant competition.

All these results confirm the usefulness of this mathematical framework, which allows accurate and reliable estimation of daily case numbers, providing a valuable complement to variant sampling in SARS-CoV-2 genomic surveillance.

These findings have several implications for public health policy and practice. First, the continuous evolution of the virus underscores the need for ongoing genomic surveillance to identify new variants of concern, as the entry of new variants may alter the epidemic situation. Second, the ability of the model to predict the spread of new variants based on their initial entry may aid in proactive planning and response. Third, our findings on variant spread in countries of different sizes can inform more nuanced, context-specific public health interventions.

Future research could focus on incorporating data from more diverse geographic locations and evaluating the model’s performance with larger, more heterogeneous datasets. In addition, further research into how factors such as population immunity and public health interventions affect variant substitution could improve our understanding of the evolutionary dynamics of SARS-CoV-2. As our understanding of the virus deepens, continued refinement of our model and others like it will remain critical to our collective efforts to combat the ongoing pandemic.

## Data Availability

All data produced are available online at GISAID and ECDC.

https://www.ecdc.europa.eu/en/publications-data/data-virus-variants-covid-19-eueea

## Acknowledgements

We gratefully acknowledge all data contributors, i.e., the Authors and their Originating laboratories responsible for obtaining the specimens, and their Submitting laboratories for generating the genetic sequence and metadata and sharing via the GISAID Initiative, on which this research is based.

We also gratefully acknowledge the ECDC authors for providing the sequence sampling data from GISAID EpiCoV and ECDC TESSy upon which this research is based.

## Footnotes

1 https://covid19.who.int/data

2 https://ec.europa.eu/eurostat/

3 https://gisaid.org/

4 https://www.ecdc.europa.eu/en

5 https://www.who.int/activities/tracking-SARS-CoV-2-variants

6 Lithuania has not reported any sequencing data from early May 2022 to late October 2022, thus it has been omitted from the Omicron variant analysis.

